# An ensemble *n*-sub-epidemic modeling framework for short-term forecasting epidemic trajectories: Application to the COVID-19 pandemic in the USA

**DOI:** 10.1101/2022.06.19.22276608

**Authors:** Gerardo Chowell, Sushma Dahal, Amna Tariq, Kimberlyn Roosa, James M. Hyman, Ruiyan Luo

## Abstract

We analyze an ensemble of *n*-sub-epidemic modeling for forecasting the trajectory of epidemics and pandemics. These ensemble modeling approaches, and models that integrate sub-epidemics to capture complex temporal dynamics, have demonstrated powerful forecasting capability. This modeling framework can characterize complex epidemic patterns, including plateaus, epidemic resurgences, and epidemic waves characterized by multiple peaks of different sizes. We systematically assess their calibration and short-term forecasting performance in short-term forecasts for the COVID-19 pandemic in the USA from late April 2020 to late February 2022. We compare their performance with two commonly used statistical ARIMA models. The best fit sub-epidemic model and three ensemble models constructed using the top-ranking sub-epidemic models consistently outperformed the ARIMA models in terms of the weighted interval score (WIS) and the coverage of the 95% prediction interval across the 10-, 20-, and 30-day short-term forecasts. In the 30-day forecasts, the average WIS ranged from 377.6 to 421.3 for the sub-epidemic models, whereas it ranged from 439.29 to 767.05 for the ARIMA models. Across 98 short-term forecasts, the ensemble model incorporating the top four ranking sub-epidemic models (Ensemble(4)) outperformed the (log) ARIMA model 66.3% of the time, and the ARIMA model 69.4% of the time in 30-day ahead forecasts in terms of the WIS. Ensemble(4) consistently yielded the best performance in terms of the metrics that account for the uncertainty of the predictions. This framework could be readily applied to investigate the spread of epidemics and pandemics beyond COVID-19, as well as other dynamic growth processes found in nature and society that would benefit from short-term predictions.

**Summary:** The COVID-19 pandemic has highlighted the urgent need to develop reliable tools to forecast the trajectory of epidemics and pandemics in near real-time. We describe and apply an ensemble *n*-sub-epidemic modeling framework for forecasting the trajectory of epidemics and pandemics. We systematically assess its calibration and short-term forecasting performance in weekly 10-30 days ahead forecasts for the COVID-19 pandemic in the USA from late April 2020 to late February 2022 and compare its performance with two different statistical ARIMA models. This framework demonstrated reliable forecasting performance and substantially outcompeted the ARIMA models. The forecasting performance was consistently best for the ensemble sub-epidemic models incorporating a higher number of top-ranking sub-epidemic models. The ensemble model incorporating the top four ranking sub-epidemic models consistently yielded the best performance, particularly in terms of the coverage rate of the 95% prediction interval and the weighted interval score. This framework can be applied to forecast other growth processes found in nature and society including the spread of information through social media.

## Introduction

The coronavirus disease 2019 (COVID-19) pandemic has amplified the critical need for reliable tools to forecast the trajectory of epidemics and pandemics in near real-time. During the early stages of the COVID-19 pandemic, multiple modeling teams embarked on the challenging task of producing short-term forecasts of the course of the COVID-19 pandemic in terms of the trajectory for the number of new cases, hospitalizations, or deaths (e.g., [1-10]). Soon after the epidemic started, our research team published short-term forecasts of the pandemic during the early outbreaks of the novel coronavirus in China [4] and subsequently focused on producing weekly forecasts for the USA [11]. In a related effort, the US COVID-19 Forecasting Hub brought together multiple research teams to synthesize weekly short-term forecasts of the COVID-19 pandemic in the USA [12]. It is time to systematically and rigorously evaluate the forecasting performance of these different pandemic forecasting efforts and document the lessons learned to continue advancing our understanding of epidemic forecasting.

Ensemble modeling approaches and models that integrate sub-epidemics to capture complex temporal dynamics have demonstrated powerful forecasting capability (e.g., [13] [14-17]). In prior work, we developed a sub-epidemic modeling framework to characterize and improve forecasting accuracy during complex epidemic waves [13]. This mathematical framework characterizes epidemic curves by aggregating multiple asynchronous sub-epidemics and outperforms simpler growth models at providing short-term forecasts of various infectious disease outbreaks [13, 18]. It is possible to model sub-epidemics associated with transmission chains that are asynchronously triggered and progress somewhat independently from the other sub-epidemics. This framework supports a family of sub-epidemic models that yield similar fits to the calibration data, but their corresponding forecasts could produce diverging trajectories.

Ensemble modeling aims to boost forecasting performance by systematically integrating the predictive accuracy tied to individual models [16, 19-21]. Past work indicates that multimodel ensemble approaches are powerful forecasting tools that frequently outperform individual models in epidemic forecasts [14, 15, 22-27]. We extend prior sub-epidemic modeling work and propose an ensemble sub-epidemic modeling framework for forecasting the trajectory of epidemics and pandemics. In this model, the sub-epidemics can start at different time points and may follow different growth rates, scaling of growth, and final sizes. Hence, this ensemble modeling framework can characterize more diverse epidemic patterns which were impossible to capture by earlier sub-epidemic models, including plateaus, epidemic resurgences, and epidemic waves characterized by multiple peaks of different sizes.

Here, we systematically assess the calibration and short-term forecasting performance in weekly 10-30 day forecasts in the context of the COVID-19 pandemic in the USA from late April 2020 to late February 2022, including the Omicron-dominated wave. We then compare the performance of the ensemble modeling framework with a set of Autoregressive Integrated Moving Average (ARIMA) models, following the EPIFORGE 2020 guidelines to report epidemic forecasts [28]. Our extended ensemble modeling framework substantially outperforms individual top-ranking sub-epidemic models and the ARIMA models based on standard performance metrics that account for the uncertainty of the predictions.

## Data

We used daily COVID-19 deaths reported in the USA from the publicly available data tracking system of the Johns Hopkins Center for Systems Science and Engineering (CSSE) from 27 February 2020 to 30 March 2022 [29]. The data is updated on the CSSE webpage once every day at 23:59 (UTC) and is read from the daily case report. The data is also publicly available in the GitHub repository [30].

### *n*-sub-epidemic model

We model epidemic trajectories comprised of one or more overlapping and asynchronous sub-epidemics. That is, the sub-epidemics are used as building blocks to characterize more complex epidemic trajectories. The mathematical equation for the sub-epidemic building block is the 3-parameter generalized-logistic growth model (GLM), which has performed well in short-term forecasts of single outbreak trajectories for different infectious diseases, including COVID-19 [31-33]. This model is given by the following differential equation:

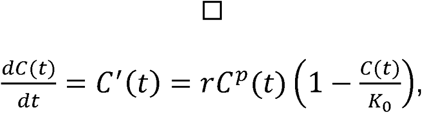

where 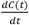 describes the curve of daily deaths over time *t*. The cumulative curve at time *t* is given by *C*(*t*), While *r* is a positive parameter denoting the growth rate per unit of time, K_0_ is the final outbreak size, and *p* ∈ [1,0] is the “scaling of growth” parameter which allows the model to capture early sub-exponential and exponential growth patterns. If *p* = 0, this equation describes a constant number of new deaths over time, while *p* = 1 indicates that the early growth phase is exponential. Intermediate values of *p* (0 < *p* < 1) describe early sub-exponential (e.g., polynomial) growth dynamics.

An *n*-sub-epidemic trajectory comprises *n* overlapping sub-epidemics and is given by the following system of coupled differential equations:

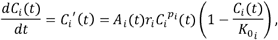

Where *C*_*i*_(*t*) tracks the cumulative number of deaths for sub-epidemic *i*, and the parameters that characterize the shape of the *i*_*th*_ sub-epidemic are given by 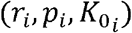, for *i* = 1, …, *n*. Thus, the 1-sub-epidemic model is equivalent to the generalized growth model described above. When *n* > 1, we model the onset timing of the (*i* + 1) _*th*_ sub-epidemic, where (*i* + 1) ≤ *n*, by employing an indicator variable given by *A*_*i*_(*t*)so that the (*i* + 1) _*th*_ sub-epidemic is triggered when the cumulative curve of the *i*_*th*_ sub-epidemic exceeds *c*_*thr*_.

The (*i* + 1) _*th*_ sub-epidemic is only triggered when 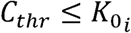. Hence, we have:

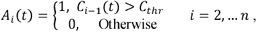

where *A*_*i*_(*t*)=1 for the first sub-epdemic. Hence, the total number of parameters that are needed to model an *n*-sub-epidemic trajectory is given by 3*n* +1. The initial number of deaths is given by *C*_1_(0) = *I*_0_, where *I*_0_ is the initial number of deaths in the observed data. The cumulative curve of the *n*-sub-epidemic trajectory is given by:

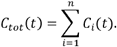

The *n*-sub-epidemic wave model can characterize diverse epidemic patterns, including epidemic plateaus where the epidemic stabilizes at a high level for an extended period, epidemic resurgences where the number of cases increases again after a low incidence period, and epidemic waves characterized by multiple peaks.

### Parameter estimation

To reduce the noise in the original data due to artificial reasons such as the weekend effects, we use the 7-day moving average of daily death series to fit the *n*-sub-epidemic model. Let

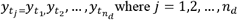

denote the smoothed daily COVID-19 death series of the epidemic trajectory based on the moving average. Here, *t*_*j*_ are the time points for the time series data, *n*_*d*_ is the number of observations, and each 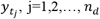, is the average of the death counts at the neighboring seven days (*t*_*j*−3_, *t*_*j*−1_, *t*_*j*−1_, *t*_*j*+1_, *t*_*j*+2_, *t*_*j*+3_). We will use this smoothed data to estimate a total of 3*n* +1 model parameters, namely 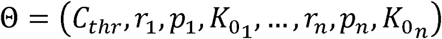. Let *f*(*t*,Θ)denote the expected curve of new COVID-19 deaths of the epidemic’s trajectory. We can estimate model parameters by fitting the model solution to the observed data via nonlinear least squares [34] or via maximum likelihood estimation assuming a specific error structure [35]. For nonlinear least squares, this is achieved by searching for the set of parameters 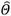 that minimizes the sum of squared differences between the observed data 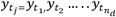 and the model mean corresponds to *f*(*t*,Θ). That is,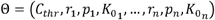 is estimated by 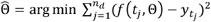.

This parameter estimation method weights each of the data points equally and does not require a specific distributional assumption for *y*_*t*_, except for the first moment *E*[*y*_*t*_] = *f*(*t*_*i*_; *Θ*). That is, the mean of the observed data at time *t* is equivalent to the expected count (e.g., number of deaths) denoted by *f*(*t*,Θ) at time *t* [36]. This method yields asymptotically unbiased point estimates regardless of any misspecification of the variance-covariance error structure. Hence, the estimated model mean 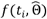 yields the best fit to observed data 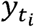 in terms of squared L2 norm. In Matlab, we can use the *fmincon* function to set the optimization problem.

To quantify parameter uncertainty, we follow a parametric bootstrapping approach which allows the computation of standard errors and related statistics in the absence of closed-form formulas [37]. We generate *B* bootstrap samples from the best-fit model 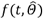, with an assumed error structure, to quantify the uncertainty of the parameter estimates and construct confidence intervals. Typically, the error structure in the data is modelled using a probability model such as the Poisson or negative binomial distribution. Because the time-series data we are fitting to involve large counts, the Poisson or negative binomial distribution can be well approximated by a normal distribution for large numbers. So, using the best-fit model 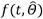, we generate *B*-times replicated simulated datasets of size *n*_*d*_, where the observation at time *t*_*j*_ is sampled from a normal distribution with mean 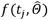 and variance 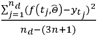. Next, we refit the model to each of the *B* simulated datasets to re-estimate parameters for each. The new parameter estimates for each realization are denoted by 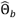 where *b* = 1,2,…,*B*. Using the sets of re-estimated parameters 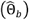, it is possible to characterize the empirical distribution of each estimate, calculate the variance, and construct confidence intervals for each parameter. The resulting uncertainty around the model fit can similarly be obtained from 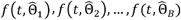.

#### Model-based forecasts with quantified uncertainty

Forecasting the model 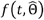, *h* days ahead provides an estimate for 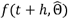. The uncertainty of the forecasted value can be obtained using the previously described parametric bootstrap method. Let

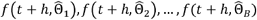

denote the forecasted value of the current state of the system propagated by a horizon of *h* time units, where 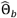 denotes the estimation of parameter set Θ from the *b*_*th*_ bootstrap sample. We can use these values to calculate the bootstrap variance as the measure of the uncertainty of the forecasts and use the 2.5% and 97.5% percentiles to construct the 95% prediction intervals (PI).

### Model selection

We considered a set of *n*-sub-epidemic models where 1≤*n* ≤2 and ranked them from best to worst according to the *AIC*_*c*_ which is given by [38, 39]:

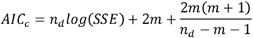

where 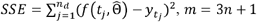 is the number of model parameters, and *n*_*d*_ is the number of data points. The *AIC*_*c*_ for the parameter estimation from the nonlinear least-squares fit, which implicitly assumes normal distribution for error.

We selected the four top ranking sub-epidemic models for further analyses. We used them to construct three ensemble sub-epidemic models, which we refer to as: Ensemble(2), Ensemble(3), and Ensemble(4). The next section describes the process of constructing these ensemble models from the top-ranking sub-epidemic models.

### Constructing Ensemble Models from top-ranking models

Ensemble models that combine the strength of multiple models may exhibit significantly enhanced predictive performance (e.g., [14-17]). Here we generate ensemble models from the weighted combination of the highest-ranking sub-epidemic models as deemed by the 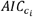 for the *i*-th model where 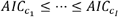 and *i* = 1, …, *I*. An ensemble derived from the top-ranking *I* models is denoted by Ensemble(*I*) and illustrated in Figure 1. Thus, Ensemble(2) and Ensemble(3) refer to the ensemble models generated from the weighted combination of the top-ranking 2 and 3 models, respectively. We compute the weight *w*_*i*_ for the *i*-th model, *i* = 1, …, *I*, where ∑*w*_*i*_ = 1 as follows:

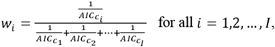

and hence *w*_*I*_ ≤ … ≤ *w*_1_.

**Figure 1.**
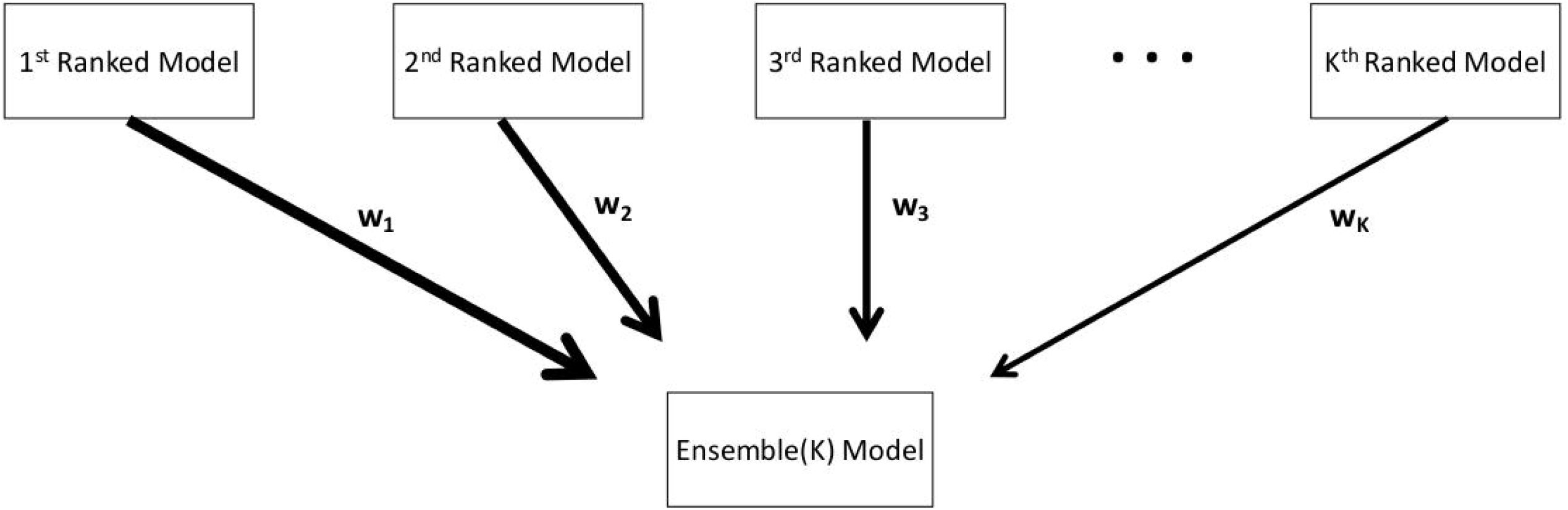
Schematic diagram of the construction of the ensemble model from the weighted combination of the highest-ranking sub-epidemic models as deemed by the 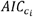 for the *i*-th model where 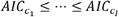 and *i* = 1, …, *I*. An ensemble derived from the top-ranking *I* models is denoted by Ensemble(I).

The estimated mean curve of daily COVID-19 deaths for the Ensemble(*I*) model is:

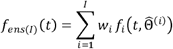

where given the training data, 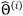 denotes the set of estimated parameters, and 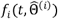 denotes the estimated mean curve of daily COVID-19 deaths, for the *i*-th model. Accordingly, we compute the weighted average and sample the bootstrap realizations of the forecasts for each model to construct the 95% CI or PI using the 2.5% and 97.5% quantiles [16]. Our MATLAB (The Mathworks, Inc) code for model fitting and forecasting is publicly available in the GitHub repository [30].

As a sensitivity analysis, we also investigated how the ensemble sub-epidemic models performed when the ensemble weights were proportional to the relative likelihood (*l*) rather than the reciprocal of the AIC_c_. Let *AIC*_*min*_ denote the minimum *AIC* from the set of models. The relative likelihood of model *i* is given by 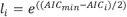 [40]. We compute the weight *w*_*i*_ for the *i*-th model where ∑*w*_*i*_ = 1 as follows:

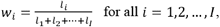

and hence *w*_*I*_ ≤ … ≤ *w*_1_.

### Auto-regressive integrated moving average models (ARIMA)

We also generated short-term predictions of the pandemic trajectory using ARIMA models to compare their performance with that of the sub-epidemic models. ARIMA models have been frequently employed to forecast trends in finance [41-43] and weather [44-46]. The ARIMA (p, d, q) process is given by

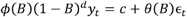

or equivalently as 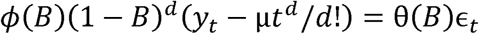, where p is the order of the AR model, d is the degree of differencing, q is the order of the MA model, {*ϵ*_*t*_} is a white noise process with mean 0 and variance σ^2^, and B denotes the backshift operator. The p-order polynomial ϕ(*z*) = 1 − ϕ_1_*z* − … − ϕ_*p*_Z^*p*^ and the q-order polynomial d θ(*z*) = 1 − θ_1_*z* − … − θ_1_*z*^*q*^ are assumed to have no roots inside the unit circle to ensure causality and invertibility. The constant C = *µ*(1 − *ϕ*_1_ − … − *ϕ*_*p*_), and µ is the mean of (1 − *B*) ^*d*^*y*_*t*_. When d=0, *µ* is the mean of *y*_*t*_.

The *auto.arima* function in the R package “forecast” is used to select orders and build the model [47]. First, the degree of differencing 0 ≤ *d* ≤ 2 is selected based on successive KPSS unit-root tests [48], which test the data for a unit root; if the test result is significant, the differenced data is tested for a unit root; and this procedure is repeated until the first insignificant result is obtained. Then given d, the orders p and q are selected based on the AIC_c_ for the d-times differenced data. For d=0 or d=1, a constant will be included if it improves the AIC_c_ value; for d>1, the constant *µ* is fixed at 0 to avoid the model having a quadratic or higher order trend, which is dangerous when forecasting. The final model is fitted using the maximum likelihood estimation.

To guarantee the forecasted values and prediction intervals are above zero, we take the following two strategies. In the first one, we conduct the ARIMA order selection and model fitting using the log-transformed data. Then we take the exponential of the forecasted values and the PI bounds to predict the incident death counts and get the PIs. We refer to this approach as the (log) ARIMA throughout the manuscript. In the second case, the negative values are set as zero. Then, it is possible that the actual coverage probability of such PIs can be smaller than the nominal value (95%). We refer to this approach as ARIMA throughout the manuscript.

### Forecasting strategy and performance metrics

We conducted short-term forecasts using the top-ranking *n*-sub-epidemic model (1 ≤ *n* ≤ 2) and three ensemble models constructed with the top-ranking sub-epidemic models namely Ensemble(2), Ensemble(3), and Ensemble(4). For comparison, we also generated short-term forecasts using the previously described ARIMA models. Overall, we conducted 588 forecasts across models.

Using a 90-day calibration period for each model, we conducted a total of 98 weekly sequential 10-day, 20-day and 30-day forecasts from 20 April 2020 to 28 February 2022, spanning five pandemic waves. This range of forecasting horizons is comparable to that investigated in prior COVID-19 forecasting studies [49]. This period covers the latter part of the early spring wave, a summer wave in 2020, a fall-winter 2020/2021 wave, the summer-fall wave in 2021, and the winter 2022 wave.

To assess the forecasting performance, we used four performance metrics: the mean absolute error (MAE), the mean squared error (MSE), the coverage of the 95% prediction intervals, and the mean interval score (MIS) [50]. The *mean absolute error* (MAE) is given by:

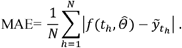

Here 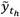 is the time series of the original death counts (unsmoothed) of the *h*-time units ahead forecasts, where *t*_*h*_ are the time points of the time series data [51]. Similarly, the *mean squared error* (MSE) is given by:

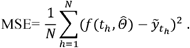

We also employed two metrics that account for prediction uncertainty: the *coverage rate of the 95% PI* e.g., the proportion of the observations that fall within the 95% PI as well as the *weighted interval score* (WIS) [50, 52] which is a proper score. The WIS and the coverage rate of the 95% PIs take into account the uncertainty of the predictions, whereas the MAE and MSE only assess the closeness of the mean trajectory of the epidemic to the observations [53].

Recent epidemic forecasting studies have embraced the Interval Score (IS) for quantifying model forecasting performance [18, 24, 49, 54]. The WIS provides quantiles of predictive forecast distribution by combining a set of ISs for probabilistic forecasts. An IS is a simple proper score that requires only a central (1−α)×100% PI [50] and is described as

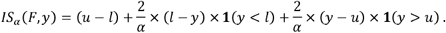

In this equation **1** refers to the indicator function, meaning that 1(*y* <, *l*) =1 if *y* < *l* and 0 otherwise. The terms *l* and *u* represent the 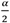 and 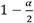 quantiles of the forecast *F*. The IS consists of three distinct quantities:

1. The sharpness of *F*, given by the width *u* − *l* of the central (1− *α*) x 100% PI.
2. A penalty term 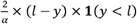 for the observations that fall below the lower end point *l* of the (1− *α*) x 100% PI. This penalty term is directly proportional to the distance between *y* and the lower end *l* of the PI. The strength of the penalty depends on the level *α*.
3. An analogous penalty term 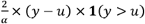 for the observations falling above the upper limit *u* of the PI.

To provide more detailed and accurate information on the entire predictive distribution, we report several central PIs at different levels (1− *α* _1_)< (1− *α*_2_) < …< (1− *α*_*K*_)along with the predictive median, *m*, which can be seen as a central prediction interval at level 1− *α* _0_ → 0.

This is referred to as the WIS, and it can be evaluated as follows:

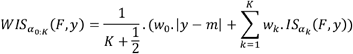

where, 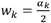for *K* = 1,2,….*K* and 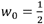. Hence, WIS can be interpreted as a measure of how close the entire distribution is to the observation in units on the scale of the observed data [10, 55].

## Results

### Quality of the sub-epidemic model fits

The best fit sub-epidemic model and three ensemble models constructed using the top-ranking sub-epidemic models (Ensemble(2), Ensemble(3), Ensemble(4)) yielded similar quality fits to 98 sequential weekly calibration periods from 20-April-2020 to 28-February-2022 (Figure 2, Table 1). For instance, the average WIS was ∼247 with little variation across models (Table 1). The coverage rate of the 95% PIs averaged 97% and ranged from 91% to 100% during the study period. Moreover, all performance metrics displayed similar temporal trends (Figure 2).

**Figure 2.**
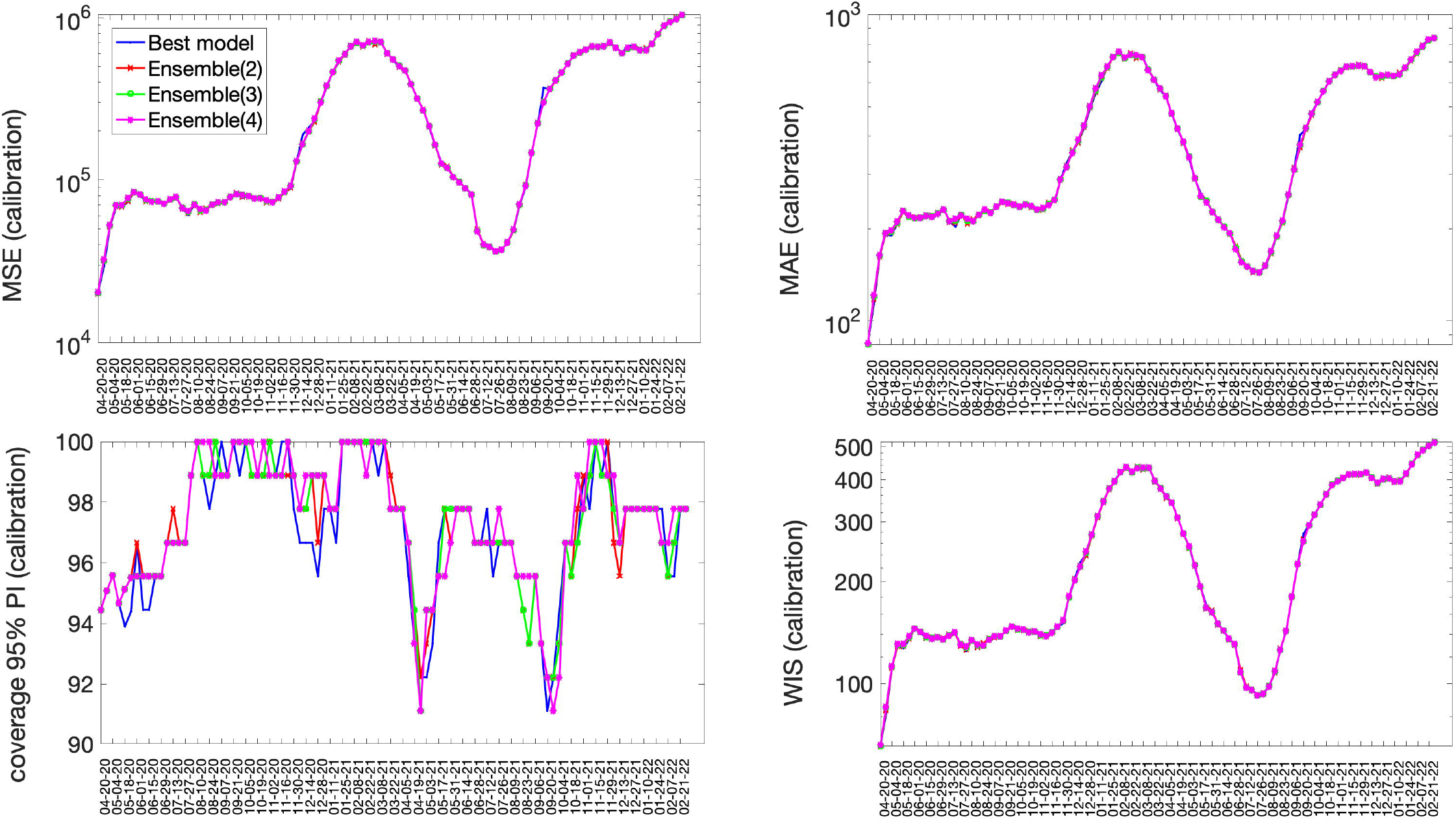
Performance metrics quantifying the quality of sub-epidemic model fits to 98 sequential weekly calibration periods of the daily time series of COVID-19 deaths in the USA from 20-April-2020 through 22-February 2022. The best fit sub-epidemic model and three ensemble models constructed using the top-ranking sub-epidemic models (Ensemble(2), Ensemble(3), Ensemble(4)) yielded similar quality fits.

**Table 1.** Mean performance metrics quantifying the quality of model fits across 98 sequential weekly calibration periods of the daily time series of COVID-19 deaths in the USA from 20-April-2020 through 22-February 2022.

| Model | Mean absolute error (MSE) | Mean squared error (MAE) | Percentage coverage of the 95% prediction interval | Weighted Interval Score (WIS) |
| --- | --- | --- | --- | --- |
| Best fit sub-epidemic model | 309260.00 | 394.74 | 97.06 | 247.28 |
| Ensemble(2) model | 308300.00 | 394.91 | 97.30 | 246.93 |
| Ensemble(3) model | 308620.00 | 395.24 | 97.46 | 247.09 |
| Ensemble(4) model | 309160.00 | 396.17 | 97.46 | 247.33 |

\*The Ensemble(*i*) model incorporates the top *i* ranked sub-epidemic models in the ensemble as described in the text.

Representative fits of the top-ranking sub-epidemic models to the daily curve of COVID-19 deaths in the USA from 27-Feb-2020 to 20-April-2020 are shown in Figure 3. Although these sub-epidemic models fit the data well, each of them results from the aggregation of two sub-epidemics characterized by different growth rates, scaling of growth, and outbreak sizes as shown in Figure 4.

**Figure 3.**
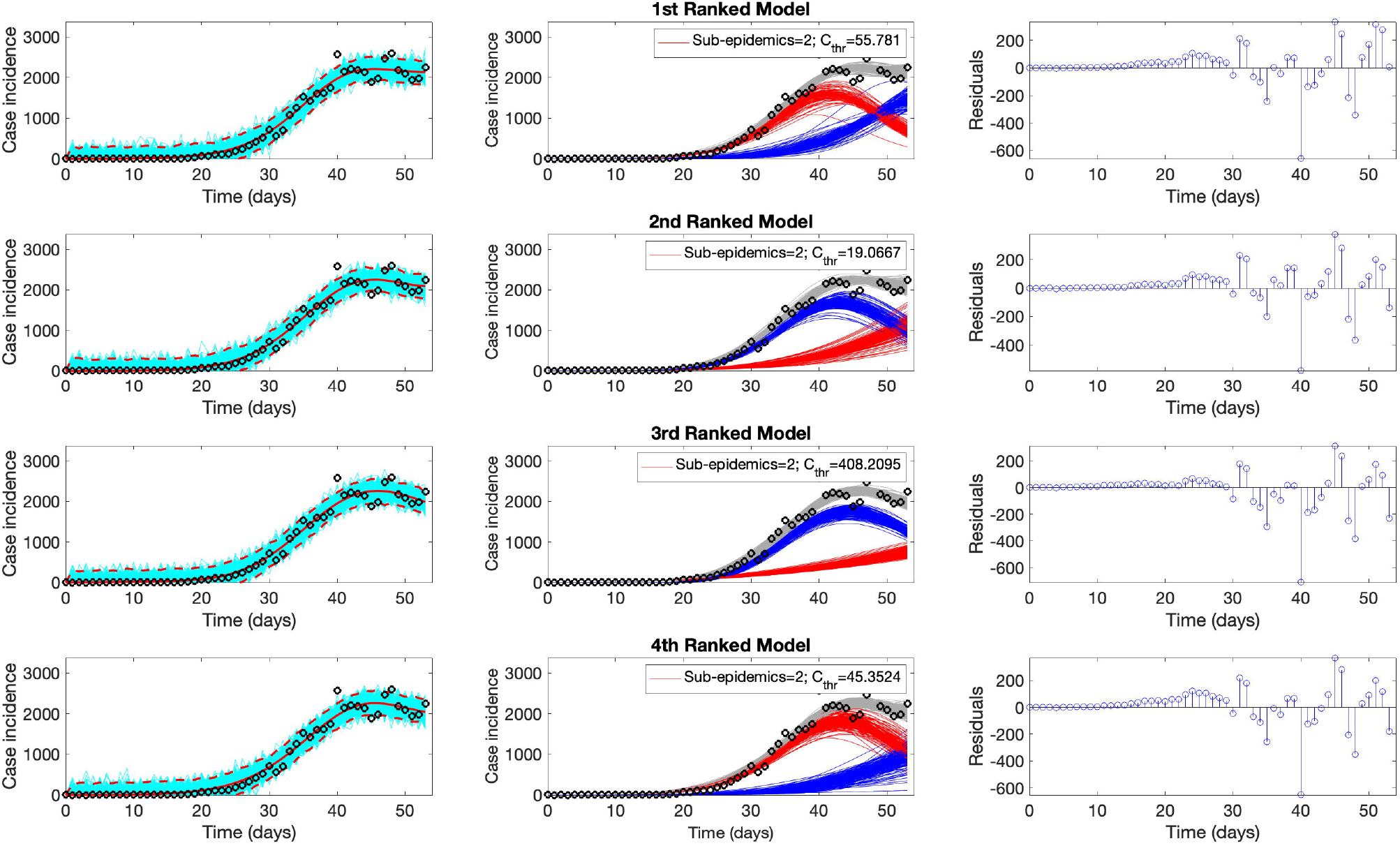
Representative fits of the top-ranking sub-epidemic models to the daily curve of COVID-19 deaths in the USA from 27-Feb-2020 to 20-April-2020. The sub-epidemic models capture well the entire epidemic curve, including the latter plateau dynamics, by considering models with two sub-epidemics. The best model fit (solid red line) and 95% prediction interval (dashed red lines) are shown in the left panels. The cyan curves correspond to the associated uncertainty from individual bootstrapped curves. The sub-epidemic profiles are shown in the center panels, where the red and blue curves represent the two sub-epidemics and the grey curves are the estimated epidemic trajectories. For each model fit, the residuals are also shown (right panels). Black circles correspond to the data points.

**Figure 4.**
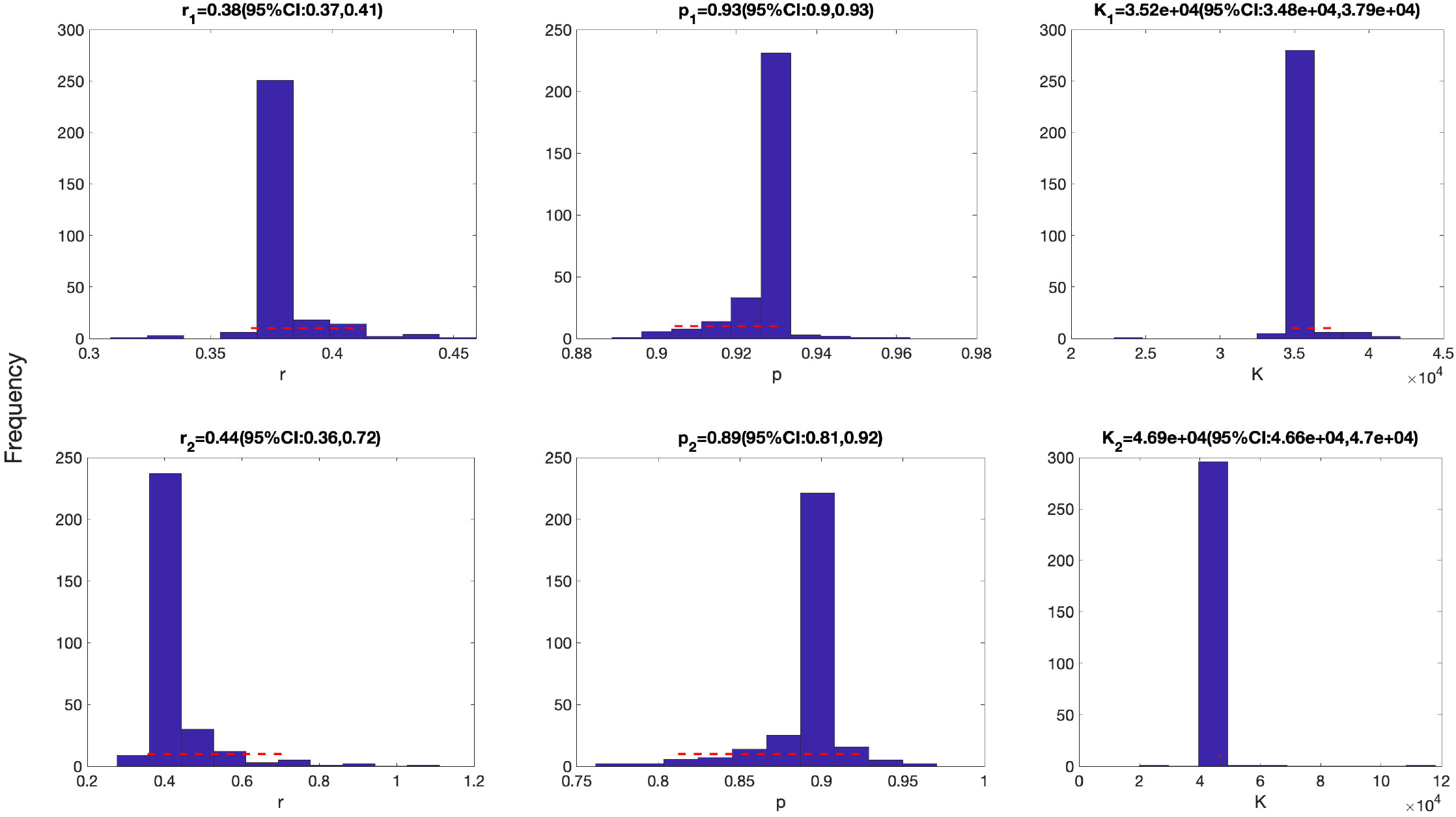
Parameter estimates for the first (top panel) and the second sub-epidemics (bottom panels) were derived for the top-ranking sub-epidemic model after fitting the sub-epidemic modeling framework to the daily curve of COVID-19 deaths in the USA from 27-Feb-2020 to 20-April-2020 (see also Figure 2). Parameter estimates for both sub-epidemics are well identified, as indicated by their relatively narrow bootstrap confidence intervals.

### Short-term forecasting performance

The best fit sub-epidemic model and three ensemble models constructed using the top-ranking sub-epidemic models (Ensemble(2), Ensemble(3), Ensemble(4)) consistently outperformed the ARIMA models in terms of the weighted interval score (WIS) and the coverage of the 95% prediction interval across the 10, 20 and 30 day short-term forecasts (Table 2). For instance, for 30-day forecasts, the average WIS ranged from 377.6 to 421.3 for the sub-epidemic models, whereas, it ranged from 439.29 to 767.05 for the ARIMA models. Across 98 short-term forecasts, the Ensemble(4) outperformed the (log) ARIMA model 66.3% of the time and the ARIMA model 69.4% of the time in 30-day ahead forecasts in terms of the WIS (Figure 5 & Figure 6). Similarly, the coverage of the 95% PI ranged from 82.2% to 88.2% for the sub-epidemic models, whereas it ranged from 58% to 60.3% for the ARIMA models in 30-day forecasts. In terms of the coverage of the 95% PI, the Ensemble(4) outperformed the (log) ARIMA model 89.8% of the time and the ARIMA model 91.8% of the time (Figure 5 & Figure Forecasting performance generally improved as the number of top-ranking sub-epidemic models included in the ensemble increased (Table 1). The Ensemble(4) model consistently yielded the best performance in terms of the metrics that account for the uncertainty of the predictions.

**Table 2.** Mean forecasting performance metrics for the sub-epidemic models (ensemble weights are proportional to the reciprocal of the AICc) and the ARIMA models across 98 sequential weekly calibration periods of the daily time series of COVID-19 deaths in the USA from 20-April-2020 through 22-February 2022. Values highlighted in bold correspond to the best performance metrics.

| Model | Mean<br>absolute<br>error (MSE) | Mean squared<br>error (MAE) | Percentage<br>coverage of the 95%<br>prediction interval | Weighted<br>Interval Score<br>(WIS) |
| --- | --- | --- | --- | --- |
| **10 days ahead** |  |  |  |  |
| Top-ranked sub-epidemic model | 551740.00 | 535.16 | 87.14 | 352.00 |
| Ensemble(2) model | 504560.00 | 516.44 | 88.88 | 331.83 |
| Ensemble(3) model | 491020.00 | 513.39 | **89.29** | 328.00 |
| Ensemble(4) model | 491740.00 | 513.14 | **89.39** | **326.56** |
| (log) ARIMA model | **424880.00** | **458.72** | 42.45 | 365.19 |
| ARIMA model | 430070.00 | 467.18 | 43.06 | 380.47 |
| **20 days ahead** |  |  |  |  |
| Top-ranked sub-epidemic model | 646880.00 | 570.34 | 85.15 | 382.90 |
| Ensemble(2) model | 576700.00 | 544.35 | 88.57 | 354.04 |
| Ensemble(3) model | 558890.00 | 540.71 | **89.59** | 350.73 |
| Ensemble(4) model | 557130.00 | 539.30 | **89.44** | **346.83** |
| (log) ARIMA model | 591980.00 | 536.22 | 51.07 | 422.41 |
| ARIMA model | **538690.00** | **528.87** | 55.05 | 404.92 |
| **30 days ahead** |  |  |  |  |
| Top-ranked sub-epidemic model | 749560.00 | 613.75 | 82.18 | 421.29 |
| Ensemble(2) model | 670740.00 | 586.52 | 87.35 | 383.36 |
| Ensemble(3) model | 650790.00 | 584.20 | **88.20** | 382.79 |
| Ensemble(4) model | **644270.00** | **579.77** | **88.16** | **377.64** |
| (log) ARIMA model | 818530.00 | 621.58 | 57.99 | 767.05 |
| ARIMA model | 656480.00 | 591.93 | 60.34 | 439.29 |

\*The Ensemble(*i*) model incorporates the top *i* ranked sub-epidemic models in the ensemble as described in the text.

**Figure 5.**
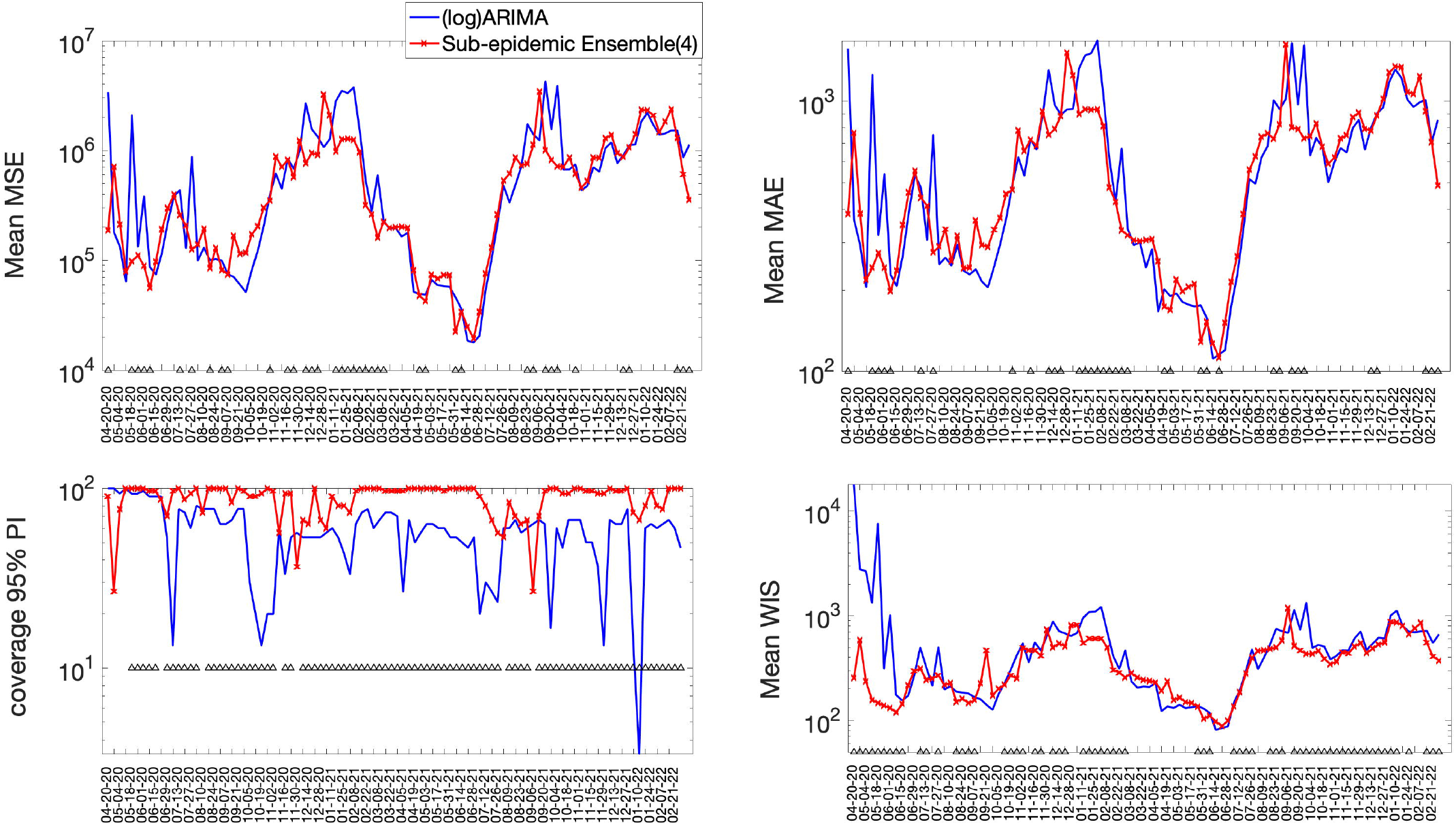
Forecasting performance metrics for the (log) ARIMA model and the Ensemble(4) model across 98 30-day forecasts. The symbol (^) indicates weekly forecasts where the Ensemble(4) model outperformed the (log) ARIMA model. For example, the Ensemble(4) outperformed the (log) ARIMA model 66.3% of the time in terms of the WIS and 89.8% of the time in terms of the coverage rate of the 95% PI (Figure 4 & Figure 6).

**Figure 6.**
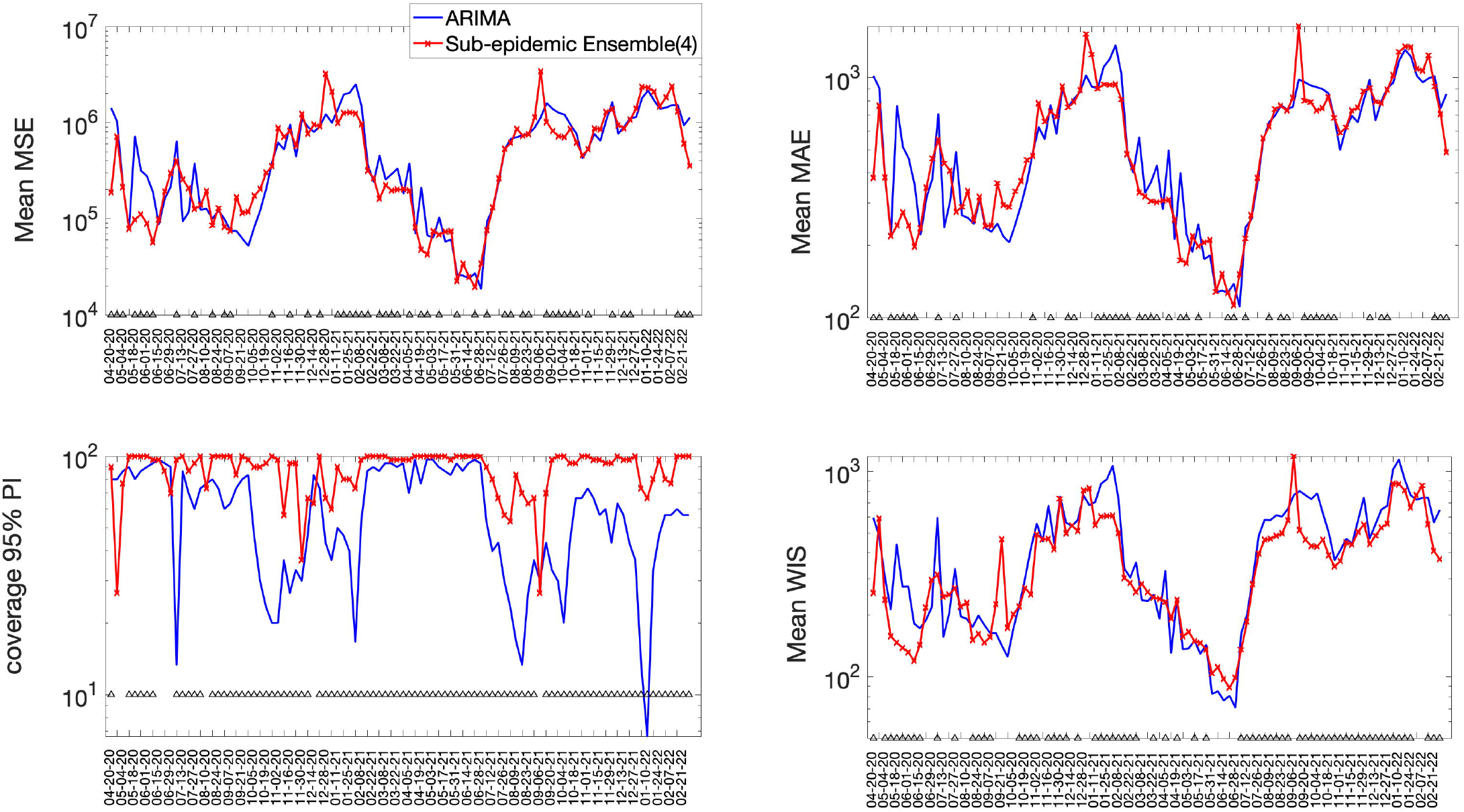
Forecasting performance metrics for the ARIMA model and the Ensemble(4) model across 98 30-day forecasts. The symbol (^) indicates weekly forecasts where the Ensemble(4) model outperforms the ARIMA model. For instance, the Ensemble(4) outperformed the ARIMA model 69.4% of the time in terms of the WIS and 91.8.8% of the time in terms of the coverage rate of the 95% PI (Figure 4 & Figure 6).

In terms of the metrics based on point estimate information, the ARIMA models showed lower overall MSE or MAE compared to the sub-epidemic models in 10 and 20-day forecasts, but the Ensemble(4) achieved the best forecasting performance in 30-day forecasts (Table 2). Overall, the forecasting performance deteriorated at longer forecasting horizons across all models considered in our study.

Representative 30-day forecasts of the top-ranking sub-epidemic models to the daily curve of COVID-19 deaths in the USA from 20-April-2020 to 20-May-2022 are shown in Figure 7. The corresponding sub-epidemic profiles of the forecasts are shown in Figure 8. These models support forecasts with diverging trajectories even though they yield similar fits to the calibration period. For instance, the top-ranked sub-epidemic model predicts a decline in the mortality curve, whereas the second-ranked model predicts a stable pattern during the next 30 days (Figure The corresponding forecasts generated from three ensemble models (Ensemble(2), Ensemble(3), Ensemble(4)) built from the top-ranking sub-epidemic models are shown in Figure 9. The individual 30-day ahead predictions across 98 forecasting periods generated by the Ensemble(4) and the ARIMA models are available in the GitHub repository [30].

**Figure 7.**
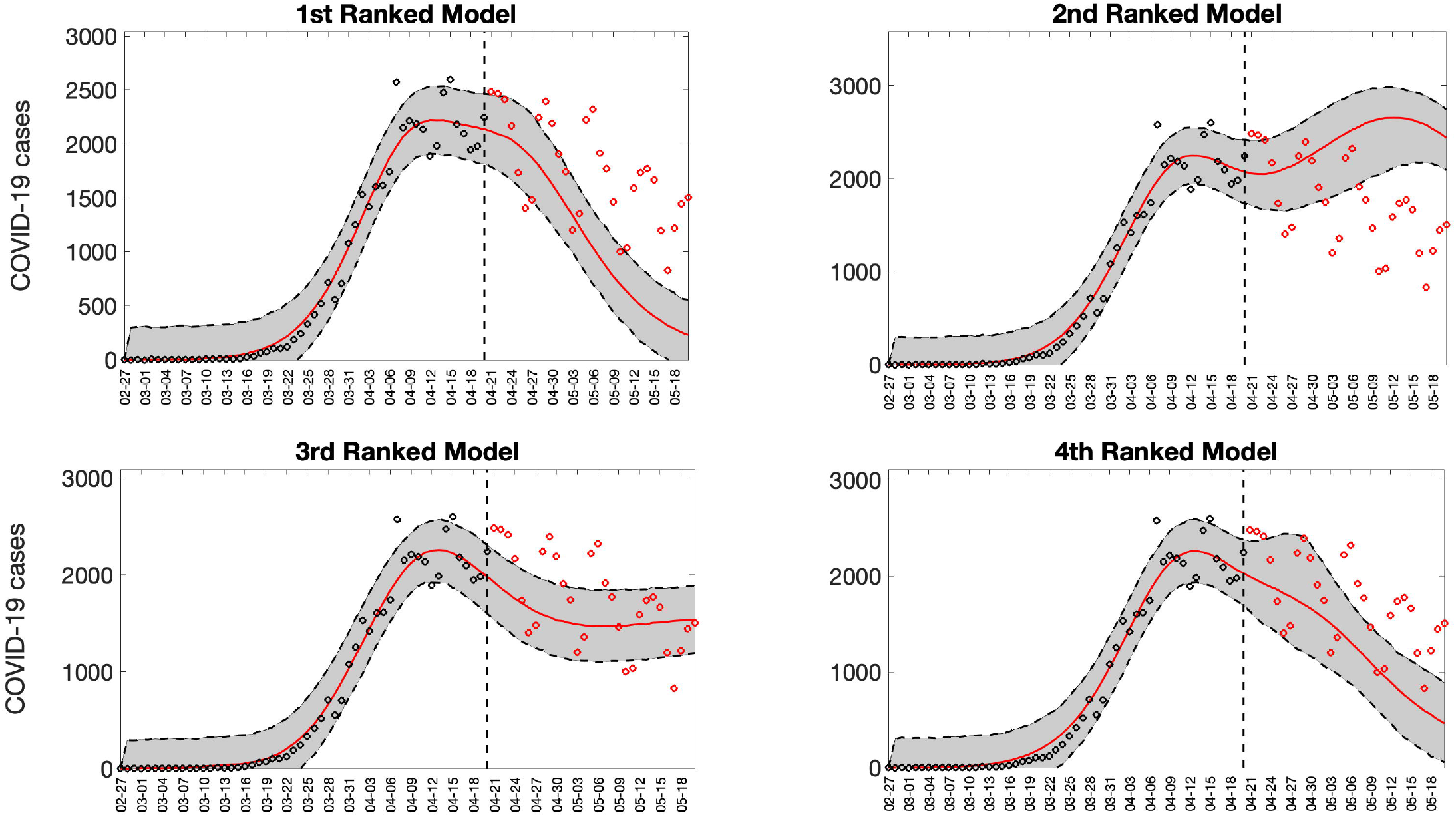
Representative 30-day forecasts of the top-ranking sub-epidemic models to the daily curve of COVID-19 deaths in the USA from 20-April-2020 to 20-May-2020. The model fit (solid line) and 95% prediction interval (shaded area) are also shown. The vertical line indicates the start time of the forecast. Circles correspond to the data points. These four top-ranking models support forecasts with diverging trajectories even though they yield similar fits to the calibration period. For instance, the 1^st^ ranked sub-epidemic model predicts a decline in the mortality curve whereas the 2^nd^ ranked model predicts a stable pattern during the next 30 days.

**Figure 8.**
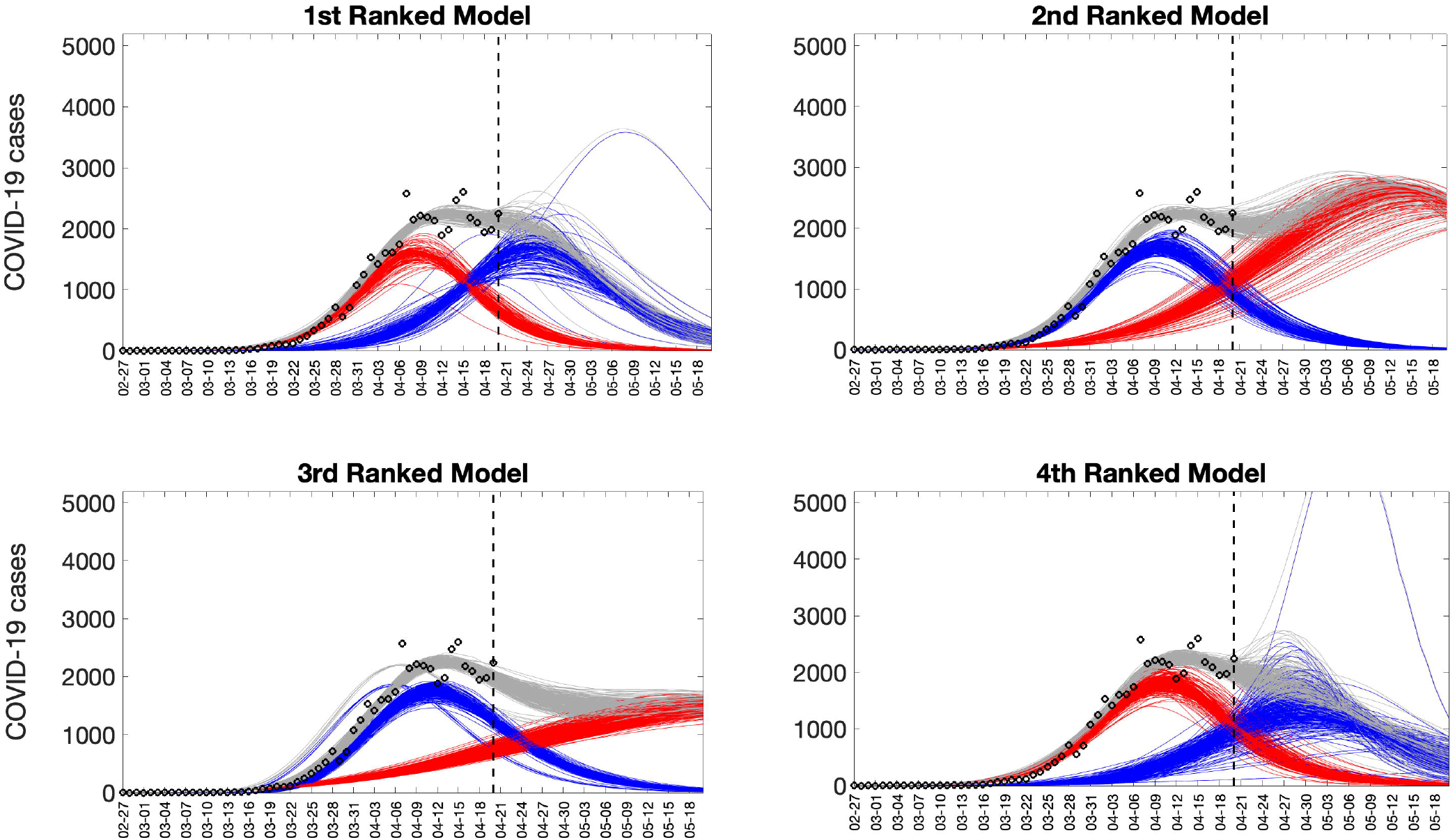
Representative sub-epidemic profiles of the forecasts derived from the top-ranking sub-epidemic models to the daily curve of COVID-19 deaths in the USA from 20-April-2020 to 20-May-2022. The model fit (solid line) and 95% prediction interval (shaded area) are also shown. Black circles correspond to the calibration data. Blue and red curves represent different sub-epidemics of the epidemic wave profile. Gray curves correspond to the overall epidemic trajectory obtained by aggregating the sub-epidemic curves. The vertical line indicates the start time of the forecast.

**Figure 9.**
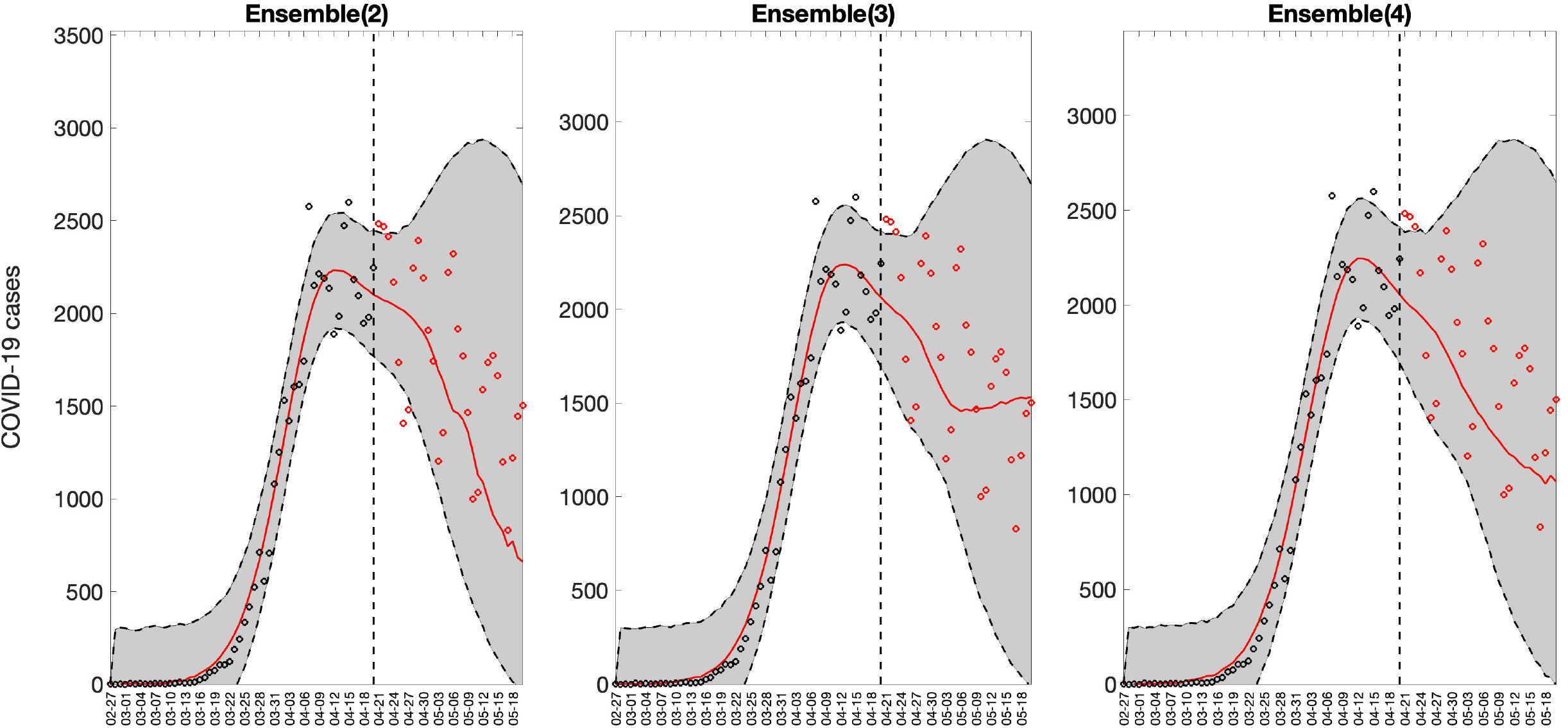
Representative sub-epidemic ensemble model forecasts (Ensemble(2), Ensemble(3), Ensemble(4)) of COVID-19 deaths in the USA from 20-April-2020 to 20-May-2022. Circles correspond to the data points. The model fits (solid line) and 95% prediction intervals (shaded area) are shown. Circles correspond to the data points. The vertical line indicates the start time of the forecast

In sensitivity analyses, defining ensemble weights as proportional to the relative likelihood did not achieve better performance relative to the ensemble models generated using weights proportional to the reciprocal of the AIC_c_. Moreover, the rank of the ensemble models was not affected by the type of weights (Table 3).

**Table 3.** Mean forecasting performance metrics for the sub-epidemic models (ensemble weights were based on the relative likelihood) and the ARIMA models across 98 sequential weekly calibration periods of the daily time series of COVID-19 deaths in the USA from 20-April-2020 through 22-February 2022. Values highlighted in bold correspond to the best performance metrics.

| Model | Mean absolute error (MSE) | Mean squared error (MAE) | Percentage coverage of the 95% prediction interval | Weighted Interval Score (WIS) |
| --- | --- | --- | --- | --- |
| **10 days ahead** |  |  |  |  |
| Top-ranked sub-epidemic model | 551740.00 | 535.16 | 87.14 | 352.00 |
| Ensemble(2) model | 548540.00 | 534.14 | **87.25** | 348.66 |
| Ensemble(3) model | 547220.00 | 533.51 | **87.25** | **347.99** |
| Ensemble(4) model | 546350.00 | 533.23 | **87.35** | **347.60** |
| (log) ARIMA model | **424880.00** | **458.72** | 42.45 | 365.19 |
| ARIMA model | 430070.00 | 467.18 | 43.06 | 380.47 |
| **20 days ahead** |  |  |  |  |
| Top-ranked sub-epidemic model | 646880.00 | 570.34 | 85.15 | 382.90 |
| Ensemble(2) model | 640240.00 | 567.90 | **85.71** | 377.27 |
| Ensemble(3) model | 640960.00 | 568.45 | **85.71** | **376.67** |
| Ensemble(4) model | 639280.00 | 567.74 | **85.56** | **376.36** |
| (log) ARIMA model | 591980.00 | 536.22 | 51.07 | 422.41 |
| ARIMA model | **538690.00** | **528.87** | 55.05 | 404.92 |
| **30 days ahead** |  |  |  |  |
| Top-ranked sub-epidemic model | 749560.00 | 613.75 | 82.18 | 421.29 |
| Ensemble(2) model | 744130.00 | 612.63 | **82.65** | **414.72** |
| Ensemble(3) model | 745230.00 | 613.21 | **82.59** | **414.54** |
| Ensemble(4) model | 743020.00 | 612.48 | **82.52** | **414.16** |
| (log) ARIMA model | 818530.00 | 621.58 | 57.99 | 767.05 |
| ARIMA model | **656480.00** | **591.93** | 60.34 | 439.29 |

## Discussion

Our ensemble sub-epidemic modeling approach outperformed individual top-ranking sub-epidemic models and a set of ARIMA models in weekly short-term forecasts covering the national trajectory of the COVID-19 pandemic in the USA from the early growth phase up until the Omicron-dominated wave. This framework has demonstrated reliable forecasting performance across different pandemic phases from the early growth phase characterized by exponential or sub-exponential growth dynamics to plateaus and new disease surges driven by the relaxation of social distancing policies or the emergence of new variants. Importantly, we found that forecasting performance consistently improved for the ensemble sub-epidemic models that incorporated a higher number of top-ranking sub-epidemic models. The ensemble model incorporating the top four ranking sub-epidemic models consistently yielded the best performance, particularly in terms of the coverage rate of the 95% prediction interval and the weighted interval score.

Our findings support the power of ensemble modeling approaches (e.g.,[14-17]). Our ensemble modeling framework derived from a family of sub-epidemic models demonstrated improved performance as the number of top-ranking sub-epidemic models included in the ensemble increased. Prior studies have documented the potential of ensemble models to enhance forecasting performance during multi-epidemic periods [14]. For instance, in the context of influenza, one study utilized “weighted density ensembles” for predicting timing and severity metrics and found that the performance of the ensemble model was comparable to that of the top individual model, albeit the ensemble’s forecasts were more stable across influenza seasons [17]. In the context of dengue in Puerto Rico, another study found that forecasts derived from Bayesian averaging ensembles outperformed a set of individual models [25]. Results from the US COVID-19 Forecasting Hub CDC were consistent with our findings in that a multimodel ensemble frequently outperformed the set of individual models.

We also evaluated short-term forecasting performance by a set of ARIMA models, as prior studies have underscored the value of ARIMA models in epidemic forecasting [56], by providing a relatively simple and transparent approach to forecasting. For instance, in the context of influenza-like-illness in the USA, a set of ARIMA models provided reasonably accurate short-term forecasts during the 2016/17 influenza season [57]. In another forecasting study during multiple seasons of influenza in the USA, an ARIMA model yielded similar short-term forecasting performance compared to other models based on the mechanistic SIR modeling framework [58]. ARIMA models have also been used for spatial prediction of the COVID-19 epidemic [59, 60]. Another study [61] showed that the ARIMA model is more effective than the Prophet time series model for forecasting COVID-19 prevalence. Finally, it is worth noting that the US COVID-19 Forecast Hub did not include an ARIMA model in its set of evaluated models [49]. Therefore, it is interesting to assess how ARIMA models perform in the context of the COVID-19 pandemic in the US.

Prior work has underscored the need to assess alternative ways of constructing ensembles from a set of individual models [14, 16]. We explored two ways of constructing the ensembles by relying on the AIC_c_ or the relative likelihood associated with the individual models. We found that the short-term forecasting performance achieved by the ensemble models was not significantly affected by the type of ensemble weights used to construct them although performance using ensemble weights based on the reciprocal of the AIC_c_ was slightly better. Further research could explore how different weighting strategies influence the forecasting performance of ensemble modeling approaches.

Short-term forecasting is an essential attribute of the models. As prior studies have underscored, longer-term forecasts are of value, but their dependability varies inversely with the time horizon. Our 20 and 30-day forecasts are most valuable for monitoring, managing, and informing the relaxing of social distancing requirements. The early detection of potential disease resurgence can signal the need for strict distancing controls, and the reports of cases can identify the geographic location of incubating sub-epidemics.

Our study is not exempt of limitations. Our analysis relied on daily time series data of COVID-19 deaths in the USA, which is inherently noisy due to heterogeneous data reporting at fine spatial scales (i.e., county-level) [62]. Noisy data complicate the ability of any mathematical model to identify meaningful signals about the impact of transmission dynamics and control interventions. To deal with the high noise levels in the data, we fitted the models to smoothed time series rather than the actual daily series, as described in the parameter estimation section. Other forecasting studies, including the US COVID-19 Forecasting Hub, have relied on weekly death counts to address this issue [49]. Beyond the COVID-19 pandemic, there is a need to establish benchmarks to systematically assess forecasting performance across a diverse catalog of mathematical models and epidemic datasets involving multiple infectious diseases, social contexts, and spatial scales.

While our analysis demonstrated the accuracy of our ensemble sub-epidemic modeling framework in forecasting the COVID-19 pandemic, the same framework could be readily used to forecast other epidemics irrespective of the type of disease and spatial scale involved. Beyond infectious diseases, this framework could also be used to forecast other biological and social growth processes, such as the epidemics of lung injury associated with e-cigarette use or vaping and the viral spread of information through social media platforms.

In summary, our ensemble sub-epidemic models provided reliable short-term forecasts of the trajectory of the COVID-19 pandemic in the USA involving multiple waves and outcompeted a set of ARIMA models. The forecasting performance of the ensemble models improved with the number of top-ranking sub-epidemic models included in the ensemble. This framework could be readily applied to investigate the spread of epidemics and pandemics beyond COVID-19 and in a range of problems in nature and society that would benefit from short-term predictions.

## Data Availability

All data referred to in this manuscript are available in our GitHub repository at https://github.com/atariq2891/An-ensemble-n-sub-epidemic-modeling-framework-for-short-term-forecasting-epidemic-trajectories

## Author contributions

Conceptualization, G.C.; methodology, G.C., J.H, R.L, validation, G.C., R.L; formal analysis, G.C., R.L; investigation, G.C., R.L; resources, G.C., ; data curation, G.C., S.D..; writing—original draft preparation, G.C., R.L; writing, review, and editing, A.T., G.C., S.D., K.R., R.L., J.M.H., ; visualization, G.C, R.L; supervision, G.C., R.L; project administration, G.C.; funding acquisition, G.C. All authors have read and agreed to the published version of the manuscript.

## References

1. Bertozzi AL, Franco E, Mohler G, Short MB, Sledge D. The challenges of modeling and forecasting the spread of COVID-19. Proc Natl Acad Sci U S A. 2020;117(29):16732–8. Epub 2020/07/04. doi: 10.1073/pnas.2006520117. PubMed PMID: 32616574; PubMed Central PMCID: PMCPMC7382213.

2. Farcomeni A, Maruotti A, Divino F, Jona-Lasinio G, Lovison G. An ensemble approach to short-term forecast of COVID-19 intensive care occupancy in Italian regions. Biometrical Journal. 2021;63(3):503–13. doi: https://doi.org/10.1002/bimj.202000189.

3. Tariq A, Undurraga EA, Laborde CC, Vogt-Geisse K, Luo R, Rothenberg R, et al. Transmission dynamics and control of COVID-19 in Chile, March-October, 2020. PLoS Neg Trop Dis. 2021;15(1):e0009070. doi: 10.1371/journal.pntd.0009070.

4. Roosa K, Lee Y, Luo R, Kirpich A, Rothenberg R, Hyman JM, et al. Real-time forecasts of the COVID-19 epidemic in China from February 5th to February 24th, 2020. Infect Dis Model. 2020;5:256–63. doi: https://doi.org/10.1016/j.idm.2020.02.002.

5. Paireau J, Andronico A, Hozé N, Layan M, Crépey P, Roumagnac A, et al. An ensemble model based on early predictors to forecast COVID-19 health care demand in France. Proceedings of the National Academy of Sciences. 2022;119(18):e2103302119. doi: doi:10.1073/pnas.2103302119.

6. Drews M, Kumar P, Singh RK, De La Sen M, Singh SS, Pandey AK, et al. Model-based ensembles: Lessons learned from retrospective analysis of COVID-19 infection forecasts across 10 countries. Science of The Total Environment. 2022;806:150639. doi: https://doi.org/10.1016/j.scitotenv.2021.150639.

7. Zhang S, Ponce J, Zhang Z, Lin G, Karniadakis G. An integrated framework for building trustworthy data-driven epidemiological models: Application to the COVID-19 outbreak in New York City. PLOS Computational Biology. 2021;17(9):e1009334. doi: 10.1371/journal.pcbi.1009334.

8. Watson GL, Xiong D, Zhang L, Zoller JA, Shamshoian J, Sundin P, et al. Pandemic velocity: Forecasting COVID-19 in the US with a machine learning & Bayesian time series compartmental model. PLOS Computational Biology. 2021;17(3):e1008837. doi: 10.1371/journal.pcbi.1008837.

9. Català M, Alonso S, Alvarez-Lacalle E, López D, Cardona P-J, Prats C. Empirical model for short-time prediction of COVID-19 spreading. PLOS Computational Biology. 2020;16(12):e1008431. doi: 10.1371/journal.pcbi.1008431.

10. Cramer EY, Ray EL, Lopez VK, Bracher J, Brennen A, Castro Rivadeneira AJ, et al. Evaluation of individual and ensemble probabilistic forecasts of COVID-19 mortality in the United States. Proc Natl Acad Sci U S A. 2022;119(15):e2113561119. Epub 2022/04/09. doi: 10.1073/pnas.2113561119. PubMed PMID: 35394862.

11. Chowell G, Tariq A, Dahal S, Roosa K. Forecasts of national COVID-19 incidence in the United States Georgia State University, School of Public Health. Epidemic Forecasting Center: GSU; 2022 [cited 2022 May 3]. Available from: https://publichealth.gsu.edu/research/coronavirus/.

12. CDC. The COVID-19 forecast hub 2021 [cited 2021 November 20]. Available from: https://covid19forecasthub.org/.

13. Chowell G, Tariq A, Hyman JM. A novel sub-epidemic modeling framework for short-term forecasting epidemic waves. BMC Med. 2019;17(1):164. doi: 10.1186/s12916-019-1406-6.

14. Chowell G, Luo R, Sun K, Roosa K, Tariq A, Viboud C. Real-time forecasting of epidemic trajectories using computational dynamic ensembles. Epidemics. 2020;30:100379. doi: https://doi.org/10.1016/j.epidem.2019.100379.

15. Viboud C, Sun K, Gaffey R, Ajelli M, Fumanelli L, Merler S, et al. The RAPIDD ebola forecasting challenge: Synthesis and lessons learnt. Epidemics. 2018;22:13–21. Epub 2017/09/30. doi: 10.1016/j.epidem.2017.08.002. PubMed PMID: 28958414; PubMed Central PMCID: PMCPMC5927600.

16. Chowell G, Luo R. Ensemble bootstrap methodology for forecasting dynamic growth processes using differential equations: application to epidemic outbreaks. BMC Medical Research Methodology. 2021;21(1):34. doi: 10.1186/s12874-021-01226-9.

17. Ray EL, Reich NG. Prediction of infectious disease epidemics via weighted density ensembles. PLoS Comput Biol. 2018;14(2):e1005910. Epub 2018/02/21. doi: 10.1371/journal.pcbi.1005910. PubMed PMID: 29462167; PubMed Central PMCID: PMCPMC5834190.

18. Tariq A, Chakhaia T, Dahal S, Ewing A, Hua X, Ofori SK, et al. An investigation of spatial-temporal patterns and predictions of the coronavirus 2019 pandemic in Colombia, 2020-2021. PLoS Negl Trop Dis. 2022;16(3):e0010228. Epub 2022/03/05. doi: 10.1371/journal.pntd.0010228. PubMed PMID: 35245285; PubMed Central PMCID: PMCPMC8926206.

19. Tebaldi C, Knutti R. The use of the multi-model ensemble in probabilistic climate projections. Philos Trans A Math Phys Eng Sci. 2007;365(1857):2053–75. Epub 2007/06/16. doi: 10.1098/rsta.2007.2076. PubMed PMID: 17569654.

20. Lindström T, Tildesley M, Webb C. A Bayesian ensemble approach for epidemiological projections. PLoS Comput Biol. 2015;11(4):e1004187. Epub 2015/05/01. doi: 10.1371/journal.pcbi.1004187. PubMed PMID: 25927892; PubMed Central PMCID: PMCPMC4415763.

21. Smith T, Ross A, Maire N, Chitnis N, Studer A, Hardy D, et al. Ensemble modeling of the likely public health impact of a pre-erythrocytic malaria vaccine. PLoS Med. 2012;9(1):e1001157. Epub 2012/01/25. doi: 10.1371/journal.pmed.1001157. PubMed PMID: 22272189; PubMed Central PMCID: PMCPMC3260300 employed by the PATH Malaria Organization which was supporting the development of RTS,S, the vaccine which is the focus of this paper. AB left PATH prior to any collaboration on this paper. All other authors have declared no competing interests. The views expressed are those of the authors.

22. McGowan CJ, Biggerstaff M, Johansson M, Apfeldorf KM, Ben-Nun M, Brooks L, et al. Collaborative efforts to forecast seasonal influenza in the United States, 2015-2016. Sci Rep. 2019;9(1):683. Epub 2019/01/27. doi: 10.1038/s41598-018-36361-9. PubMed PMID: 30679458; PubMed Central PMCID: PMCPMC6346105 consulting for S.K. Analytics. The remaining authors declare no competing interests.

23. Johansson MA, Apfeldorf KM, Dobson S, Devita J, Buczak AL, Baugher B, et al. An open challenge to advance probabilistic forecasting for dengue epidemics. Proc Natl Acad Sci U S A. 2019;116(48):24268–74. Epub 2019/11/13. doi: 10.1073/pnas.1909865116. PubMed PMID: 31712420; PubMed Central PMCID: PMCPMC6883829.

24. Roosa K, Tariq A, Yan P, Hyman JM, Chowell G. Multi-model forecasts of the ongoing Ebola epidemic in the Democratic Republic of Congo, March 2013-October 2019. J R Soc Interface. 2020;17(169):20200447. doi: doi:10.1098/rsif.2020.0447.

25. Yamana TK, Kandula S, Shaman J. Superensemble forecasts of dengue outbreaks. J R Soc Interface. 2016;13(123). Epub 2016/10/14. doi: 10.1098/rsif.2016.0410. PubMed PMID: 27733698; PubMed Central PMCID: PMCPMC5095208.

26. Novaes de Amorim A, Deardon R, Saini V. A stacked ensemble method for forecasting influenza-like illness visit volumes at emergency departments. PLOS ONE. 2021;16(3):e0241725. doi: 10.1371/journal.pone.0241725.

27. Kim J-S, Kavak H, Züfle A, Anderson T. COVID-19 ensemble models using representative clustering. SIGSPATIAL Special. 2020;12(2):33–41. doi: 10.1145/3431843.3431848.

28. Pollett S, Johansson MA, Reich NG, Brett-Major D, Del Valle SY, Venkatramanan S, et al. Recommended reporting items for epidemic forecasting and prediction research: The EPIFORGE 2020 guidelines. PLOS Medicine. 2021;18(10):e1003793. doi: 10.1371/journal.pmed.1003793.

29. CSSE Covid-19 Timeseries [Internet]. 2022 [cited May 20, 2022]. Available from: https://github.com/CSSEGISandData/COVID-19/blob/master/csse_covid_19_data/csse_covid_19_time_series/time_series_covid19_confirmed_US.csv.

30. n-subepidemic ensemble modeling framework [Internet]. 2022. Available from: https://github.com/atariq2891/An-ensemble-n-sub-epidemic-modeling-framework-for-short-term-forecasting-epidemic-trajectories

31. Shanafelt DW, Jones G, Lima M, Perrings C, Chowell G. Forecasting the 2001 Foot-and-Mouth Disease Epidemic in the UK. Ecohealth. 2017. Epub 2017/12/15. doi: 10.1007/s10393-017-1293-2. PubMed PMID: 29238900.

32. Chowell G, Hincapie-Palacio D, Ospina J, Pell B, Tariq A, Dahal S, et al. Using Phenomenological Models to Characterize Transmissibility and Forecast Patterns and Final Burden of Zika Epidemics. PLoS Curr. 2016;8. Epub 2016/07/02. doi: 10.1371/currents.outbreaks.f14b2217c902f453d9320a43a35b9583. PubMed PMID: 27366586; PubMed Central PMCID: PMCPMC4922743.

33. Pell B, Kuang Y, Viboud C, Chowell G. Using phenomenological models for forecasting the 2015 Ebola challenge. Epidemics. 2018;22:62–70. Epub 2016/12/04. doi: 10.1016/j.epidem.2016.11.002. PubMed PMID: 27913131.

34. Banks HT, Hu S, Thompson WC. Modeling and inverse problems in the presence of uncertainty: CRC Press; 2014.

35. Roosa K, Luo R, Chowell G. Comparative assessment of parameter estimation methods in the presence of overdispersion: a simulation study. Math Biosci Eng. 2019;16(5):4299–313. Epub 2019/09/11. doi: 10.3934/mbe.2019214. PubMed PMID: 31499663.

36. Myung IJ. Tutorial on maximum likelihood estimation. Journal of Mathematical Pyschology; 2003. p. 90–100.

37. Friedman J, Hastie T, Tibshirani R. The Elements of Statistical Learning : Data Mining, Inference, and Prediction. New York, NY.: Springer-Verlag New York; 2009.

38. Sugiura N. Further analysts of the data by akaike’ s information criterion and the finite corrections. Communications in Statistics-theory and Methods. 1978;7:13–26.

39. Hurvich CM, Tsai C-L. Regression and time series model selection in small samples. Biometrika. 1989;76:297–307.

40. Burnham KP, Anderson DR. Model selection and multimodel inference: a practical information-theoretic approach. 2 ed: Springer-Verlag, New York, NY; 2002. p. 488.

41. Prapanna M, Shit L, Goswami. S. Study of effectiveness of time series modeling (ARIMA) in forecasting stock prices. International Journal of Computer Science, Engineering and Applications. 2014;4.2(13).

42. Adebiyi AA, Adewumii A, Ayo C. Stock price prediction using the ARIMA model. UKSim-AMSS 16th International Conference on Computer Modelling and Simulation: IEEE; 2014.

43. Almasarweh M, Alwadi S. ARIMA model in predicting banking stock market data. Modern Applied Science 2018;12(11):309.

44. Tektaş M. Weather Forecasting Using ANFIS and ARIMA MODELS. Environmental Research, Engineering and Management. 2010;51(1):5–10.

45. Shamsnia SA, Shahidi N, Liaghat A, Sarraf A, Vahdat SF. Modeling of weather parameters using stochastic methods (ARIMA model)(case study: Abadeh Region, Iran). International conference on environment and industrial innovation 2011.

46. Dimri T, Ahmad S, Sharif M. Time series analysis of climate variables using seasonal ARIMA approach. Journal of Earth System Science. 2020;129(1):149. doi: 10.1007/s12040-020-01408-x.

47. Hyndman RJ, Khandakar Y. Automatic Time Series Forecasting: The forecast Package for R. Journal of Statistical Software. 2008;27(3):1–22. doi: 10.18637/jss.v027.i03.

48. Kwiatkowski D, Phillips PCB, Schmidt P, Shin Y. Testing the null hypothesis of stationarity against the alternative of a unit root: How sure are we that economic time series have a unit root? Journal of Econometrics. 1992;54(1):159–78. doi: https://doi.org/10.1016/0304-4076(92)90104-Y.

49. Bracher J, Ray EL, Gneiting T, Reich NG. Evaluating epidemic forecasts in an interval format. PLoS Comput Biol. 2021;17(2):e1008618. doi: 10.1371/journal.pcbi.1008618.

50. Gneiting T, Raftery AE. Strictly Proper Scoring Rules, Prediction, and Estimation. Journal of the American Statistical Association. 2007;102(477):359–78. doi: 10.1198/016214506000001437.

51. Kuhn M, Johnson K. Applied predictive modeling: New York: Springer; 2013.

52. M4Competition. Competitor’s Guide: Prizes and Rules. 2018. Available from: https://www.m4.unic.ac.cy/wp-content/uploads/2018/03/M4-Competitors-Guide.pdf.

53. Funk S, Camacho A, Kucharski AJ, Lowe R, Eggo RM, Edmunds WJ. Assessing the performance of real-time epidemic forecasts: A case study of Ebola in the Western Area region of Sierra Leone, 2014-15. PLoS Comput Biol. 2019;15(2):e1006785. Epub 2019/02/12. doi: 10.1371/journal.pcbi.1006785. PubMed PMID: 30742608.

54. Hwang E. Prediction intervals of the COVID-19 cases by HAR models with growth rates and vaccination rates in top eight affected countries: Bootstrap improvement. Chaos Solitons Fractals. 2022;155:111789–. Epub 2022/01/03. doi: 10.1016/j.chaos.2021.111789. PubMed PMID: 35002103.

55. Bracher J, Ray EL, Gneiting T, Reich NG. Evaluating epidemic forecasts in an interval format. PLoS Comput Biol. 2021;17(2):e1008618. doi: 10.1371/journal.pcbi.1008618.

56. Rguibi MA, Moussa N, Madani A, Aaroud A, Zine-Dine K. Forecasting Covid-19 Transmission with ARIMA and LSTM Techniques in Morocco. SN Comput Sci. 2022;3(2):133–. Epub 2022/01/14. doi: 10.1007/s42979-022-01019-x. PubMed PMID: 35043096.

57. Kandula S, Shaman J. Near-term forecasts of influenza-like illness: An evaluation of autoregressive time series approaches. Epidemics. 2019;27:41–51. doi: https://doi.org/10.1016/j.epidem.2019.01.002.

58. Reich NG, Brooks LC, Fox SJ, Kandula S, McGowan CJ, Moore E, et al. A collaborative multiyear, multimodel assessment of seasonal influenza forecasting in the United States. Proc Natl Acad Sci U S A. 2019;116(8):3146–54. Epub 2019/01/17. doi: 10.1073/pnas.1812594116. PubMed PMID: 30647115; PubMed Central PMCID: PMCPMC6386665.

59. Roy S, Bhunia GS, Shit PK. Spatial prediction of COVID-19 epidemic using ARIMA techniques in India. Model Earth Syst Environ. 2021;7(2):1385–91. Epub 2020/08/25. doi: 10.1007/s40808-020-00890-y. PubMed PMID: 32838022; PubMed Central PMCID: PMCPMC7363688.

60. Jacques Demongeot KO, Mustapha Rachdi, Lahoucine Hobbad, Mohamed Alahiane, Siham Iggui, Jean Gaudart, Idir Ouassou,. he application of ARIMA model to analyze COVID-19 incidence pattern in several countries. J Math Comput Sci. 2021;12.

61. Naresh Kumara SS. COVID-19 pandemic prediction using time series forecasting models. 11th International Conference on Computing, Communication and Networking Technologies (ICCCNT): IEEE; 2020.

62. Taylor KS, Taylor JW. Interval forecasts of weekly incident and cumulative COVID-19 mortality in the United States: A comparison of combining methods. PLOS ONE. 2022;17(3):e0266096. doi: 10.1371/journal.pone.0266096.

